# COVID-19 infections post-vaccination by HIV status in the United States

**DOI:** 10.1101/2021.12.02.21267182

**Authors:** Sally B. Coburn, Elizabeth Humes, Raynell Lang, Cameron Stewart, Brenna C Hogan, Kelly A. Gebo, Sonia Napravnik, Jessie K. Edwards, Lindsay E. Browne, Lesley S. Park, Amy C. Justice, Kirsha Gordon, Michael A. Horberg, Julia M. Certa, Eric Watson, Celeena R Jefferson, Michael Silverberg, Jacek Skarbinski, Wendy A Leyden, Carolyn F. Williams, Keri N. Althoff, for the Corona-Infectious-Virus Epidemiology Team (CIVETs) of the NA-ACCORD of IeDEA

## Abstract

**Importance:** Recommendations for additional doses of COVID vaccine are restricted to people with HIV who have advanced disease or unsuppressed HIV viral load. Understanding SARS-CoV-2 infection risk post-vaccination among PWH is essential for informing vaccination guidelines.

**Objective:** Estimate the risk of breakthrough infections among fully vaccinated people with (PWH) and without (PWoH) HIV in the US.

**Design, setting, and participants:** The Corona-Infectious-Virus Epidemiology Team (CIVET)-II cohort collaboration consists of 4 longitudinal cohorts from integrated health systems and academic health centers. Each cohort identified individuals ≥18 years old, in-care, and fully vaccinated for COVID-19 through 30 June 2021. PWH were matched to PWoH on date fully vaccinated, age group, race/ethnicity, and sex at birth. Incidence rates per 1,000 person-years and cumulative incidence of breakthrough infections with 95% confidence intervals ([,]) were estimated by HIV status. Cox proportional hazards models estimated adjusted hazard ratios (aHR) of breakthrough infections by HIV status adjusting for demographic factors, prior COVID-19 illness, vaccine type (BNT162b2, [Pfizer], mRNA-1273 [Moderna], Jansen Ad26.COV2.S [J&J]), calendar time, and cohort. Risk factors for breakthroughs among PWH, were also investigated.

**Exposure:** HIV infection

**Outcome:** COVID-19 breakthrough infections, defined as laboratory evidence of SARS-CoV-2 infection or COVID-19 diagnosis after an individual was fully vaccinated.

**Results:** Among 109,599 individuals (31,840 PWH and 77,759 PWoH), the rate of breakthrough infections was higher in PWH versus PWoH: 44 [41, 48] vs. 31 [29, 33] per 1,000 person-years. Cumulative incidence at 210 days after date fully vaccinated was low, albeit higher in PWH versus PWoH overall (2.8% versus 2.1%, log-rank p<0.001, risk difference=0.7% [0.4%, 1.0%]) and within each vaccine type. Breakthrough infection risk was 41% higher in PWH versus PWoH (aHR=1.41 [1.28, 1.56]). Among PWH, younger age (18-24 versus 45-54), history of COVID-19 prior to fully vaccinated date, and J&J vaccination (versus Pfizer) were associated with increased risk of breakthroughs. There was no association of breakthrough with HIV viral load suppression or CD4 count among PWH.

**Conclusions and Relevance:** COVID-19 vaccination is effective against infection with SARS-CoV-2 strains circulating through 30 Sept 2021. PWH have an increased risk of breakthrough infections compared to PWoH. Recommendations for additional vaccine doses should be expanded to all PWH.

## INTRODUCTION

As vaccines for COVID-19 continue to be distributed in the United States (US), breakthrough infections are occurring in a small percentage of vaccinated individuals, observed in both clinical trials^1–3^ and observational settings.^4–9^ Characterizing breakthrough infections is critical for efforts to curb the pandemic and to deepen our understanding of the scope of population immunity conferred by vaccination. Additionally, the rate of breakthrough infections is necessary information for setting public health policies, including prioritization of additional doses for the primary series and boosters.

Immunocompromised individuals may be particularly at risk for breakthrough infections relative to the US general population. Studies have indicated that solid-organ transplant recipients are at increased risk of breakthrough infections relative to individuals who are not immunocompromised.^10–12^ People with HIV (PWH) are an immunocompromised population who may benefit from additional vaccination doses to minimize their risk of acquiring SARS-CoV-2 infection, though this has not been comprehensively investigated. Clinical trial data for vaccines available in the US were insufficiently powered to stratify outcomes by HIV status.^1–3^ Although two observational studies have compared the risk of breakthrough infection among PWH versus without HIV (PWoH), these were limited in geographic scope, number of individuals with HIV, and generalizability to the broader population of PWH in the US.^10,13^ These studies also did not address differences in breakthrough rates among PWH by vaccine type or by clinically relevant factors including HIV viral suppression or CD4 count at the time of vaccination, which could impact risk.

The US Centers for Disease Control and Prevention’s (CDC’s) guidance for PWH is specific to those with advanced or untreated HIV and includes the recommendation of an additional primary dose 28 days after a second dose of the mRNA-1273 (Moderna) or BNT162 (Pfizer) vaccines and eligibility for a subsequent booster dose; there is no recommendation for an additional primary series does of Janssen Ad26.COV2.S (J&J), however, a booster (of any vaccine type) is recommended 2 months after a single J&J dose.^14^ Current recommendations do not consider treated PWH who are HIV suppressed, many of whom have chronic immune impairment such as partially recovered CD4 counts, or persistent immune activation and dysfunction from their HIV infection.^15^ These PWH may still be at increased risk for breakthrough infections compared to PWoH and may benefit from an additional dose in their mRNA primary series or a booster 2 months after a single dose of J&J. More detailed and generalizable evaluations of breakthrough infections among PWH versus PWoH are needed to inform US vaccine guidelines on the primary vaccine schedule for all PWH.

Our objective was to determine if HIV status is associated with an increased rate or risk of breakthrough infection among fully vaccinated individuals in a collaboration of four longitudinal cohorts from integrated health systems or academic health centers in the US, by vaccination type, and by immune/viral suppression status at the time of vaccination among PWH.

## METHODS

### Study population

The Corona-Infectious-Virus Epidemiology Team (CIVET)-II cohort, established in September 2021, is comprised of four cohorts from integrated health systems and academic health centers that contribute longitudinal data on PWH to the North American AIDS Cohort Collaboration on Research and Design (NA-ACCORD).^16^ A collaboration allowed for a large and geographically diverse sample of PWH from which inferences could be made. The cohorts span several US geographic regions and include the following: Kaiser Permanente Mid-Atlantic States (Maryland, District of Columbia, northern Virginia), Kaiser Permanente Northern California, University of North Carolina Chapel Hill HIV Clinic, and the Veterans Aging Cohort Study (VACS) which is a sample of all PWH receiving care within the National US Veterans Affairs Healthcare System. The four cohorts received approval from their local institutional review boards (IRB) and the overall project was approved by the Johns Hopkins Bloomberg School of Public Health IRB.

The CIVET-II cohort collaboration participants include PWH and PWoH observed during the COVID-19 pandemic. Adults (≥18 years old) “in-care” (determined by unique criteria for each site; see **Supplement Table S1**) and fully vaccinated against COVID-19 between December 11, 2020 (date of Emergency Use Authorization of the first COVID-19 vaccine) and June 30, 2021 were eligible. Full vaccination status was defined using CDC criteria depending on the vaccine type: a) 14 days after the second dose for those receiving Pfizer or Moderna mRNA vaccines; or b) 14 days after the single dose of the J&J viral vector vaccine.^17^ Individuals were excluded from the study population if they received a vaccine that was not authorized in the US.

Each fully vaccinated PWH was matched to three PWoH on the date considered fully vaccinated (+/- 14 days), 10-year age group (18-24, 25-34, 35-44, 45-54, 55-64, 65-74, ≥75 years), race/ethnicity (Black/African American, white, Hispanic, Asian, other, unknown), and sex at birth (female or male). When necessary, PWH could be matched to individuals either one age group above or below their category. If three matches were not available, PWH could be matched to one or two PWoH to maximize the study population size. All cohorts completed this matching schema except for VACS (N=65,440), which has its own long-standing schema to match each veteran with HIV to two veterans without HIV on age, race/ethnicity, sex, and clinical site at the establishment of the cohort and during its dynamic enrollment as veterans enter HIV care; the VACS participants were not matched on vaccination date.^18^ All variables, irrespective of HIV status, including matching factors described above, were abstracted from electronic medical records.

### Outcome: Breakthrough infection after fully vaccinated

The first infection with SARS-CoV-2 or COVID-19 illness diagnosed after the date an individual is fully vaccinated (14 days after the last required dose) was defined as a breakthrough case (**Supplemental Figure S1**). Incident COVID-19 cases were identified using: 1) positive or detectable SARS-CoV-2 nucleic acid amplification assay (NAAT) or antigen test; and/or 2) International Statistical Classification of Diseases and Related Health Problems (ICD)-10 codes U07.1 (specific to COVID), B34.2 (Coronavirus infection, unspecified), B97.21 (SARS-associated coronavirus causing disease classified elsewhere), B97.29 (other coronavirus as the cause of diseases classified elsewhere, or J12.81 (pneumonia due to SARS-associated coronavirus).

For each patient, all positive SARS-CoV-2 laboratory tests were identified. Any additional positive laboratory tests and/or ICD-10 diagnoses occurring within +/- 90 days of that date of a positive or detectable SARS-CoV-2 test result were considered persistent infection (**Supplemental Figure S1**). The 90-day window was implemented based on the CDC’s suggestion that diagnosis of re-infections should not be considered until 90-days after evidence of initial infection as test positivity and symptoms can be prolonged.^19^ In instances where there was a COVID-19 diagnosis within the 90-day window of a positive or detectable test result, only the date of the laboratory test was used to define the date of the breakthrough case; positive laboratory results were prioritized over diagnosis codes for calculation of breakthrough date due to their greater specificity. This same process was completed for ICD-10 diagnoses that did not occur within +/- 90 window of another laboratory test.

### Exposure: HIV infection

PWH were identified using HIV registries or ICD diagnosis codes for HIV, depending on the participating cohort (see **Supplemental Table S1**). PWoH were classified as such if there was no evidence of infection using these same sources as of December 11, 2020 (including no positive ELISA or Western blot tests, no HIV RNA measurements, no ICD diagnosis codes, and not found in an HIV registry).

### Covariates

In addition to demographics used to match fully vaccinated PWH to PWoH (i.e., age, race and ethnicity, sex at birth), covariates of interest included the type of primary series vaccine (Pfizer, Moderna, J&J), vaccine dose received ≥90 days after the completion of the primary series, and evidence of COVID-19 infection prior to date fully vaccinated (history of COVID-19). COVID-19 diagnoses prior to date fully vaccinated included both infection prior to any vaccination and those that occurred in the window between the first dose but before full vaccination (partial breakthrough). These were identified using the approach used to identify our main outcome.

Among PWH, CD4 count and HIV-1 RNA viral suppression status were collected as close to date full vaccination as possible (within a window of 1 Jan 2020 to full vaccination) and at antiretroviral therapy (ART) initiation (within a window of 12 months prior to 1 month after). HIV viral suppression was defined as <50 copies/mL, which was the highest lower limit of quantification used across the health systems. History of AIDS diagnosis (clinical diagnosis^20^ or CD4 count <200 cells/mm^3^, depending on the cohort) prior to date fully vaccinated was included.

### Statistical analysis

Study entry for eligible individuals was the date they were fully vaccinated (defined as 14 days after final dose of series). Individuals were followed to the date of breakthrough infection, death, disenrollment from the health system (applicable to only 2 of the 4 health systems), 210 days (7 months) post-fully vaccinated date, or until September 30^th^, 2021, whichever occurred first. We assessed the distributions of demographic and clinical characteristics to determine potential differences between PWH and PWoH. Differences were detected using χ^2^ tests for categorical variables.

Incidence rates and 95% confidence intervals ([,]) of COVID-19 breakthrough infections after the date fully vaccinated were calculated per 1,000 person-years (PY) for each month overall, by HIV status, and by vaccine type.

The 7-month (210 days) cumulative incidence of breakthrough infections by HIV status was estimated from the date fully vaccinated. Cumulative incidence estimates were stratified by HIV status, and among PWH, CD4 count (<200, 200-349, 350-499, and 500 cells/mm^3^) and viral suppression status (<50 copies/mL). Cumulative incidence was also estimated by HIV status for each vaccine type. Log-rank tests were calculated to test for significant differences in cumulative incidence on Kaplan-Meier curves.

We compared the risk of breakthrough infection by HIV status using a Cox proportional hazard models to estimate unadjusted and adjusted hazard ratios (aHR) with 95% confidence intervals. Adjustment factors included: sex, race/ethnicity, age, primary vaccine series type, cohort, and an interaction between a history of COVID-19 and 3-month calendar period (January-March, April-June, and July-September 2021). The interaction term was included because Schoenfeld’s residuals suggested the hazard of breakthrough was not proportional for those with a history of COVID-19 during our study period [p<0.01], and a likelihood ratio test suggested a better fit of the model with the interaction [p<0.01]. Subgroup analyses included: a) excluding those with a history of COVID-19 prior to full vaccination; b) excluding VACS participants due to the differences in matching strategies.

Among PWH, HIV-associated risk factors for breakthrough infections were examined, including CD4 count (<200 cells/mm^3^, 200-349 cells/mm^3^, 350-499 cells/mm^3^, and ≥500 cells/mm^3^), and detectable viral load (<50 copies/mL), accounting for the covariates included in the main analysis of PWH and PWoH. Sub-group analyses excluding those with a history of COVID-19 prior to full vaccination and VACS participants were conducted.

All analyses were conducted in R and a p-value<0.05 was considered statistically significant.

## RESULTS

Of the 109,719 fully vaccinated patients, 120 (40 PWH and 80 PWoH) were excluded due to mixing of vaccine type within the primary series, resulting in a study population of 109,599 patients (31,840 PWH and 77,759 PWoH) (**Table 1**). Most patients were 55 years and older (71%), male (92%), and non-Hispanic Black (41%). Most participants received either the Pfizer (51%) or the Moderna (43%) vaccines. A small minority received J&J (6%). Although we did not match on vaccine type, the distribution of vaccine type by HIV status did not differ by more than 2 percentage points. Twenty-six percent of PWH received an additional COVID-19 vaccine dose after their primary series compared to 12% of PWoH. Differences in the characteristics of PWH who did and did not receive an additional vaccine dose after their primary series can be found in **Supplementary Table S2**). Among PWH, 15% had a history of AIDS prior to vaccination. At the time of full vaccination, 88% of PWH were virally suppressed and the median CD4 count was 622 cells/mm^3^ (IQR: 424, 846).

**Table 1:** Characteristics at date SARS-CoV-2 fully vaccinated of people with (PWH) and without HIV (PWoH), N=109,599

| Characteristic | Overall, N = 109,599<sup>1</sup> | PWoH, N = 77,759<sup>1</sup> | PWH, N = 31,840<sup>1</sup> |
| --- | --- | --- | --- |
| **Age (years)** |  |  |  |
| 18-24 | 293 (0.3%) | 212 (0.3%) | 81 (0.3%) |
| 25-34 | 4,273 (3.9%) | 2,754 (3.5%) | 1,519 (4.8%) |
| 35-44 | 9,827 (9.0%) | 6,595 (8.5%) | 3,232 (10.2%) |
| 45-54 | 17,791 (16.2%) | 12,275 (15.8%) | 5,516 (17.3%) |
| 55-64 | 35,529 (32.4%) | 24,907 (32.0%) | 10,622 (33.4%) |
| 65-74 | 31,981 (29.2%) | 23,631 (30.4%) | 8,350 (26.2%) |
| 75+ | 9,905 (9.0%) | 7,385 (9.5%) | 2,520 (7.9%) |
| **Sex** |  |  |  |
| Male | 101,187 (92.3%) | 71,551 (92.0%) | 29,636 (93.1%) |
| Female | 8,412 (7.7%) | 6,208 (8.0%) | 2,204 (6.9%) |
| **Ethnicity and Race** |  |  |  |
| Non-Hispanic white | 42,005 (38.3%) | 29,374 (37.8%) | 12,631 (39.7%) |
| Non-Hispanic Black/African American | 44,487 (40.6%) | 31,635 (40.7%) | 12,852 (40.4%) |
| Hispanic | 14,707 (13.4%) | 10,709 (13.8%) | 3,998 (12.6%) |
| Non-Hispanic Asian | 3,857 (3.5%) | 2,837 (3.6%) | 1,020 (3.2%) |
| Other | 3,432 (3.1%) | 2,392 (3.1%) | 1,040 (3.3%) |
| Unknown | 1,111 (1.0%) | 812 (1.0%) | 299 (0.9%) |
| **Month fully vaccinated** |  |  |  |
| January 2021 | 1,169 (1.1%) | 826 (1.1%) | 343 (1.1%) |
| February 2021 | 11,603 (10.6%) | 7,963 (10.2%) | 3,640 (11.4%) |
| March 2021 | 28,775 (26.3%) | 20,293 (26.1%) | 8,482 (26.6%) |
| April 2021 | 41,680 (38.0%) | 29,770 (38.3%) | 11,910 (37.4%) |
| May 2021 | 19,793 (18.1%) | 14,288 (18.4%) | 5,505 (17.3%) |
| June 2021 | 6,579 (6.0%) | 4,619 (5.9%) | 1,960 (6.2%) |
| **Primary vaccination series type** |  |  |  |
| Pfizer | 55,906 (51.0%) | 39,186 (50.4%) | 16,720 (52.5%) |
| Moderna | 46,529 (42.5%) | 33,355 (42.9%) | 13,174 (41.4%) |
| J&J | 7,164 (6.5%) | 5,218 (6.7%) | 1,946 (6.1%) |
| **Additional dose after primary series** | 17,334 (15.8%) | 9,071 (11.7%) | 8,263 (26.0%) |
| **COVID prior to fully vaccinated** | 6,563 (6.0%) | 4,417 (5.7%) | 2,146 (6.7%) |
| **CD4 at ART initiation (cells/mm<sup>3</sup>)** |  |  | 366.00 (200.00, 580.00) |
| Unknown |  |  | 14,530 |
| **AIDS before fully vaccinated** |  |  | 1,644 (14.8%) |
| Unknown |  |  | 20,738 |
| **CD4 at fully vaccinated (cells/mm<sup>3</sup>)** |  |  | 622.00 (424.00, 846.00) |
| Unknown |  |  | 6,090 |
| **Suppressed HIV RNA at fully vaccinated (<50 copies/mL)** |  |  | 24,500 (88.1%) |
| Unknown |  |  | 4,031 |

<sup>1</sup>n (%) except for CD4 counts where the medians (interquartile ranges) are reported
P-values for all demographic characteristics were statistically significantly different comparing PWH versus PWoH using $\alpha=0.05$ as the threshold

The overall incidence rate of breakthrough infections was 35 [33, 36] per 1,000 person-years (1,767 breakthroughs among 50,731 PY). The incidence rate of breakthrough infections was higher in PWH (44 [41, 48] per 1,000 PY) versus PWoH (31 [29, 33] per 1,000 PY). The incidence rate of breakthrough infections was highest with the J&J vaccine (59 [51, 69] per 1,000 PY), followed by Pfizer (40 [37, 42] per 1,000 PY), and Moderna (26 [24, 28] per 1,000 PY, **Supplemental Table S3**). Stratified by vaccine type, the rate of breakthroughs was consistently higher among PWH versus PWoH. The higher rate of breakthroughs among PWH remained after stratifying by calendar time, especially in July and August of 2021 (**Figure 1**). There was a bimodal distribution of breakthrough infections by calendar month which was congruent with waves of the US epidemic occurring in January-February 2021 and July-September 2021.

**Figure 1:**
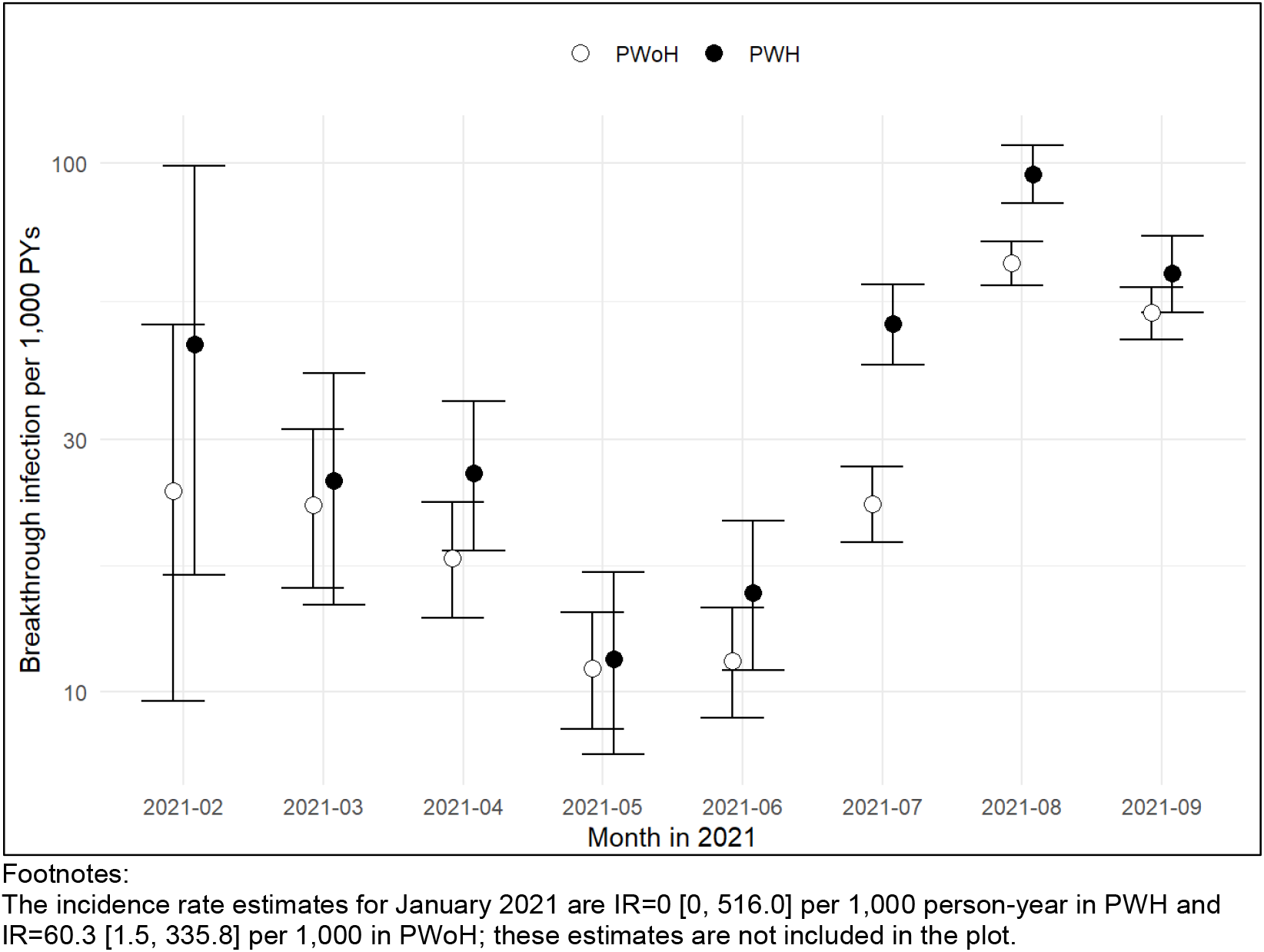
Trends in SARS-CoV-2 vaccine breakthrough incidence rates and 95% confidence intervals, by HIV status

The cumulative incidence of breakthrough infections regardless of vaccine type at 210 days (i.e., 7 months) after date fully vaccinated was 2.3% [2.2%, 2.5%] (**Figure 2a**), and higher among PWH (2.8% [2.6%, 3.1%]) versus PWoH (2.1% [2.0% 2.3%], log rank p<0.01). The risk difference between the cumulative incidence of breakthrough infections was 0.68% (95% CI 0.38, 0.98) higher in PWH. When stratified by CD4 count, PWH with lower CD4 counts at full vaccination had higher cumulative incidence of breakthroughs, although this was not statistically significant (log-rank p=0.18 after excluding PWoH, **Figure 2b**). Similarly, PWH with unsuppressed HIV viral load had a higher risk of breakthrough infection those with suppressed viral load, but this was not statistically significant (log-rank p=0.47 after excluding PWoH, **Figure 2c**). PWH had higher cumulative incidence of breakthrough, regardless of CD4 count or HIV viral load suppression, as compared to PWoH (**Figures 2b and 2c**).

**Figure 2:**
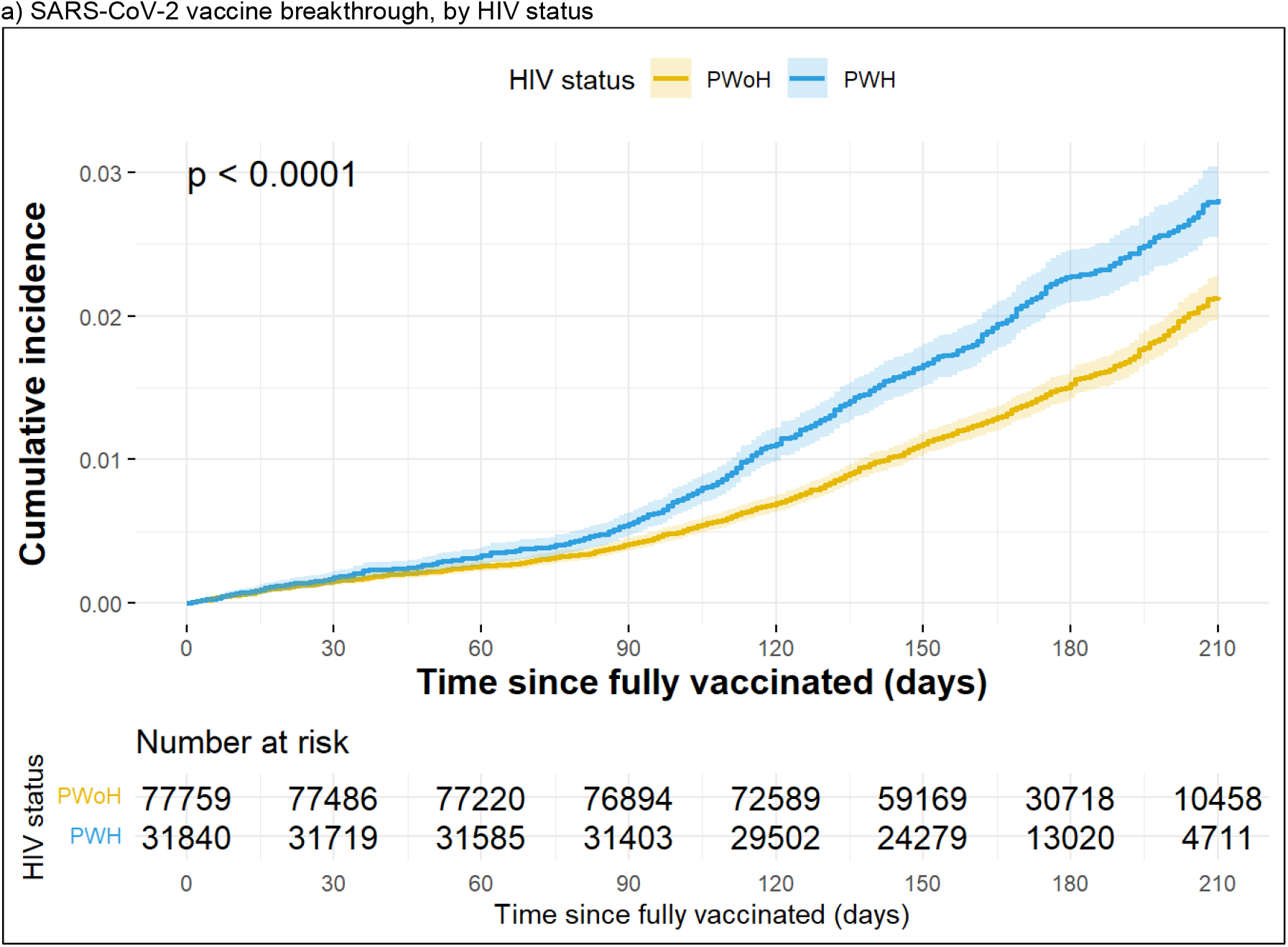
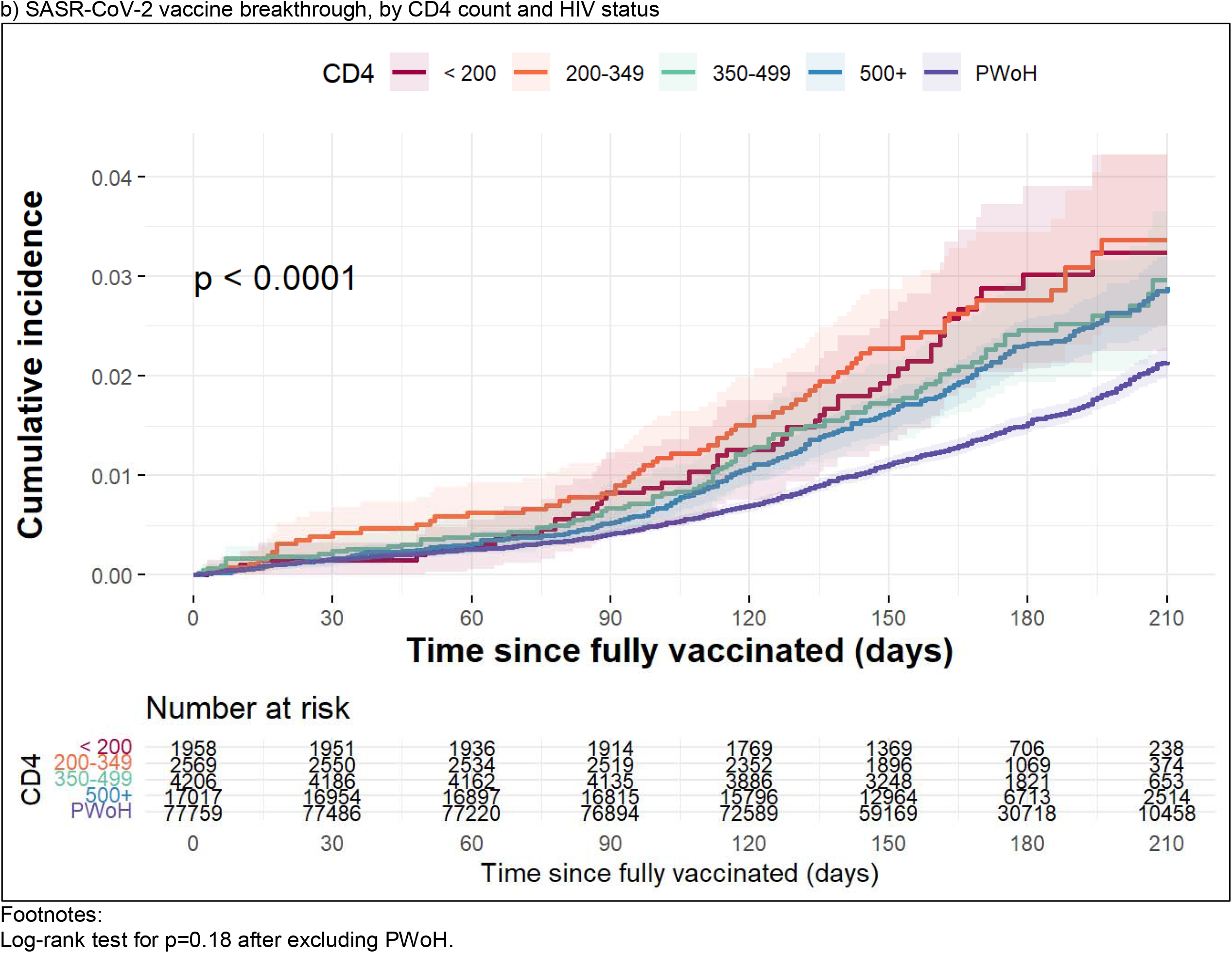
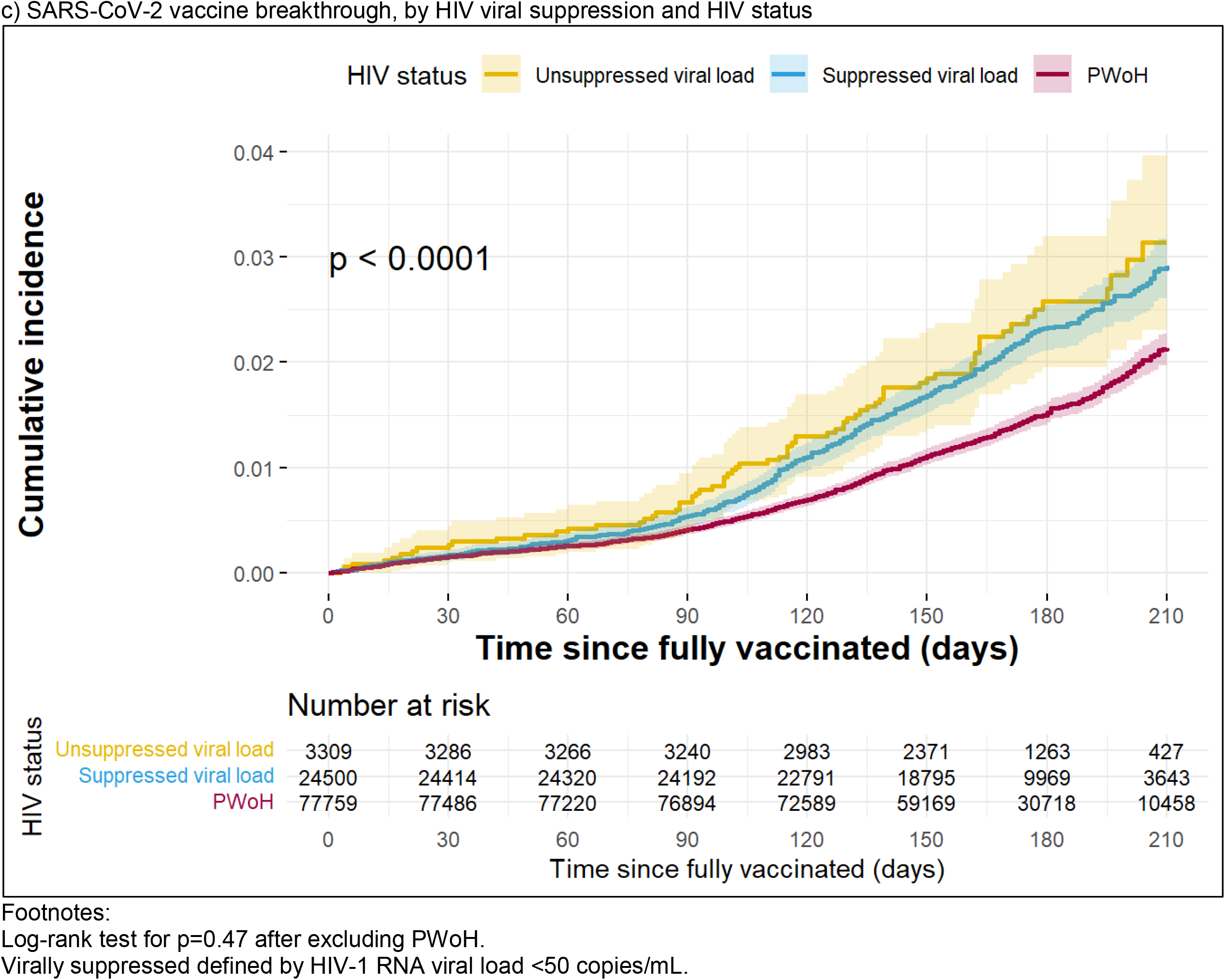
Cumulative incidence of SARS-CoV-2 vaccine breakthrough (and 95% confidence intervals represented by the shading), stratified by a) HIV status, b) CD4 count and HIV status, and c) HIV viral suppression and HIV status

The risk of breakthrough infection differed by vaccine type (**Figure 3**). The overall risk of breakthrough was highest with J&J (3.3% [2.7, 3.8%]), followed by Pfizer (2.6% [2.5%, 2.8%]) and Moderna (1.7% [1.6%, 1.9%]) at 210 days post-full vaccination. The risk remained consistently higher among PWH versus PWoH across vaccine types.

**Figure 3:**
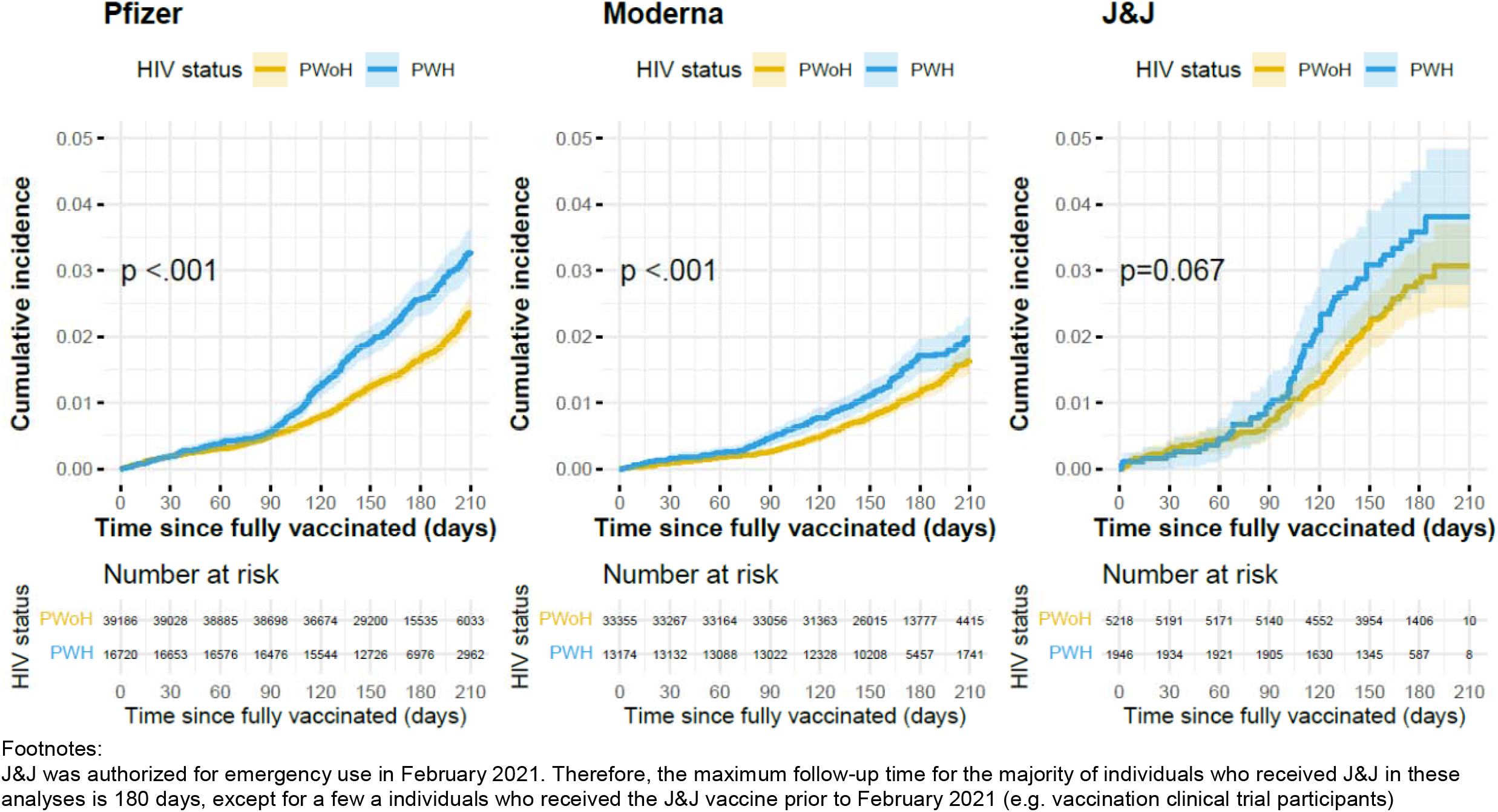
Cumulative incidence of SARS-CoV-2 vaccine breakthrough infection (and 95% confidence intervals represented by the shading), stratified by HIV status and primary vaccination series type

PWH had a significantly higher risk of breakthrough infection compared to PWoH (aHR=1.41 [1.28, 1.56]) after adjusting for covariates of interest (described above) (**Table 2**). The association was robust in subgroup analyses where: a) individuals with of history of COVID-19 were excluded (103,036 individuals after exclusion; aHR=1.40 [1.26, 1.56]); and b) VACS individuals were excluded (44,159 individuals after exclusion; aHR=1.54 [1.33, 1.79]).

**Table 2:** Crude and adjusted hazard ratios and 95% confidence intervals SARS-CoV-2 vaccination breakthrough infections

|  | HR | 95% CI | aHR | 95% CI |
| --- | --- | --- | --- | --- |
| **Among PWH and PWoH (N=109,599)<sup>a</sup>** |  |  |  |  |
| HIV status |  |  |  |  |
| PWoH | 1.00 | -- | 1.00 | -- |
| PWH | **1.43** | **1.30, 1.58** | **1.41** | **1.28, 1.56** |
| **Among PWH (N=25,478)<sup>b</sup>** |  |  |  |  |
| Sex |  |  |  |  |
| Male | 1.00 |  | 1.00 |  |
| Female | 0.98 | 0.70, 1.37 | 0.96 | 0.67, 1.36 |
| Ethnicity and Race |  |  |  |  |
| Non-Hispanic white | 1.00 |  | 1.00 |  |
| Non-Hispanic Black/African American | 1.08 | 0.89, 1.31 | 1.04 | 0.85, 1.27 |
| Hispanic | **1.32** | **1.02, 1.70** | 1.05 | 0.81, 1.37 |
| Non-Hispanic Asian | 1.32 | 0.84, 2.07 | 1.02 | 0.64, 1.62 |
| Other | 0.98 | 0.59, 1.63 | 0.91 | 0.55, 1.52 |
| Unknown | 1.31 | 0.58, 2.95 | 1.01 | 0.44, 2.29 |
| Age (years) |  |  |  |  |
| 18-24 | **3.21** | **1.31, 7.86** | **2.94** | **1.19, 7.24** |
| 25-34 | 1.09 | 0.74, 1.60 | 0.98 | 0.66, 1.46 |
| 35-44 | 1.06 | 0.79, 1.42 | 1.00 | 0.74, 1.34 |
| 45-54 | 1.00 |  | 1.00 |  |
| 55-64 | **0.73** | **0.58, 0.93** | **0.76** | **0.60, 0.96** |
| 65-74 | **0.63** | **0.49, 0.81** | **0.73** | **0.56, 0.95** |
| 75+ | **0.63** | **0.44, 0.91** | 0.76 | 0.52, 1.11 |
| Primary vaccination series type |  |  |  |  |
| Moderna | **0.61** | **0.51, 0.74** | **0.61** | **0.51, 0.74** |
| Pfizer | 1.00 |  | 1.00 |  |
| J&J | **1.56** | **1.17, 2.09** | **1.35** | **1.01, 1.82** |
| COVID-19 prior to fully vaccinated |  |  |  |  |
| No | 1.00 |  | 1.00 |  |
| Yes | **3.08** | **2.49, 3.83** | **2.87** | **2.30, 3.56** |
| Calendar period |  |  |  |  |
| Jan – March | 1.57 | 0.92, 2.69 | 1.68 | 0.99, 2.88 |
| April – June | 1.00 |  | 1.00 |  |
| July – Sept | **3.22** | **2.34, 4.44** | **2.92** | **2.12, 4.03** |
| HIV RNA |  |  |  |  |
| Unsuppressed (≥50 copies/mL) | 1.00 |  | 1.00 |  |
| Suppressed (<50 copies/mL) | 0.97 | 0.75, 1.26 | 1.04 | 0.80, 1.35 |
| CD4 count at fully vaccinated (cells/mm<sup>3</sup>) |  |  |  |  |
| <200 | 1.00 |  | 1.00 |  |
| 200-349 | 1.03 | 0.71, 1.50 | 1.02 | 0.70, 1.48 |
| 350-499 | 0.84 | 0.59, 1.19 | 0.83 | 0.59, 1.19 |
| ≥500 | 0.79 | 0.59, 1.07 | 0.74 | 0.54, 1.01 |

Abbreviations: HR=crude hazard ratio. aHR=adjusted hazard ratio. 95% CI=95% confidence interval.
<sup>a</sup>Adjusted for age, sex, race and ethnicity, primary vaccination series type, COVID-19 prior to fully vaccinated, 3-month calendar period, an interaction of COVID-19 prior to fully vaccinated and 3-month calendar period, and cohort.
<sup>b</sup>N=6,362 (20% of all PWH) were excluded due to missing CD4 or HIV RNA measurements. Adjusted for the covariates in the table and cohort.

Among PWH (25,478 after exclusion for missing risk factor data), older age (55-74 years) was associated with decreased risk of breakthrough, and younger age (18-24 years) was associated with increased risk, as compared to individuals ages 44-54 (**Table 2**). Compared to those with Pfizer, individuals who received the Moderna primary vaccination series had a reduced risk of breakthrough (aHR: 0.61 [0.51, 0.74]), while those with the J&J primary vaccination series had an increased risk of breakthrough (aHR: 1.35 [1.01, 1.82]). The risk of breakthrough infection was higher during the Delta variant (B.1.617.2) surge in the July-September 2021 3-month calendar period relative to April-June 2021 (reference period). There was no association with unsuppressed (vs. suppressed) HIV viral load and the risk or breakthrough decreased with increasing CD4 count, but this was not statistically significant. There was a near three-fold increase in the risk of breakthrough among those with evidence of a history of COVID-19 (aHR: 2.87 [2.30, 3.56]). After removing those with a history of COVID-19 prior to the date fully vaccinated (23,630 individuals after exclusion), the estimated associations of age, vaccine type, 3-month calendar period, unsuppressed viral load, and CD4 count were similar (data not shown). After removing VACS participants (9,517 individuals after exclusion), the risk of breakthrough among those with a history of COVID-19 prior to the date fully vaccinated was attenuated to a null association (aHR=0.93 [0.56, 1.56]) and the estimated associations of age, vaccine type, 3-month calendar period, unsuppressed viral load, and CD4 count were similar (data not shown).

## DISCUSSION

Among 109,599 fully vaccinated individuals receiving care at four academic or integrated health care systems across varied geographic regions in the US, breakthroughs were uncommon in vaccinated PWH and PWoH. As anticipated, only 2.3% of vaccinated individuals had breakthrough infections in the 7 months after being fully vaccinated, further demonstrating the effectiveness of the vaccines against the SARS-CoV-2 variants circulating prior to 30 Sept 2021. However, there was a consistently higher occurrence of breakthrough infections among PWH (compared to PWoH) following full COVID-19 vaccination. The higher risk of breakthrough infection among PWH versus PWoH persisted in regression analyses after adjustment for demographic factors and other covariates of interest. The cumulative incidence 210 days post full vaccination was higher in PWH compared with PWoH (2.8% vs. 2.1%). The higher rates and risk remained when stratified by vaccine type, though breakthrough occurrence overall was highest in J&J and lowest in Moderna vaccine preparations. Among PWH, the cumulative incidence and relative risks of breakthroughs was not statistically significantly different by CD4 count or HIV viral load suppression. Even among PWH with higher CD4 counts and suppressed HIV viral loads, the risk of breakthrough was greater than in PWoH, suggesting the CDC’s recommendations for an additional dose in the primary mRNA vaccination series and a booster after a single dose of J&J should not be restricted to PWH who have advanced disease or unsuppressed HIV viral load.

This is a time-to-event investigation of breakthrough infections by HIV status in a large study population followed longitudinally across several geographic regions in the US. Two prior observational studies found no significant association between HIV status and breakthrough infection risk.^10,13^ In the present analysis, we found a 41% increased risk of breakthrough infection in PWH versus PWoH after adjusting for demographic factors. This discrepancy may be due to differences in sample size and/or calendar periods of follow-up. Our findings are consistent with studies that implicate other immunocompromising conditions (e.g., solid-organ transplant, use of immune suppressing medications, active cancer diagnosis) have increased risk of breakthrough.^10–12^ Regardless of CD4 count, the cumulative incidence of breakthroughs was higher among PWH versus PWoH, which is suggestive of residual immune function abnormalities despite CD4 count recovery.

Our observation of differential breakthrough risk by vaccine type is consistent with studies showing lower effectiveness for J&J relative to the mRNA vaccines,^21^ and, among mRNA vaccines, more breakthroughs among those with Pfizer primary series than Moderna (though estimates were not always statistically significant).^10,13,22^

Among PWH, the finding that older age was associated with lower risk of breakthrough infections are likely not representative of a biological association, but rather behavioral modifications by older individuals to follow prevention guidelines more closely.^23,24^ The observation that breakthrough risk varied by calendar period was expected, given the state of the pandemic during those months, which aligns with the surge of infections seen with the Delta variant.^25^ The association we observed between history of COVID-19 prior to vaccination and increased breakthrough risk among PWH may be a reflection of increased exposure and/or adoption (or lack thereof) of prevention measures. For example, PWH with increased exposure (perhaps occupational) prior to being fully vaccinated may have had persistent increased exposure post-full vaccination, leading to increased breakthroughs. This may also reflect the increased burden of underlying comorbidities among people aging with HIV that increased their vulnerability to COVID-19. Detecting COVID-19 prior to, and after, being fully vaccinated may also be a function of lower barriers to accessing care and regularly seek care. These hypotheses are supported by the attenuation (to null) of the association of a history of COVID-19 prior to fully vaccinated with breakthrough in the subgroup analysis excluding VACS participants.

Our findings are not necessarily reflective of all PWH in the US, as we were only able to assess individuals with access to care. We may not have captured those who had less regular access to health care, who may also be at greater risk for infection. For instance, one study showed that individuals with substance use disorders have a higher risk of breakthrough infections.^22^ Individuals engaged in HIV care may have more health-seeking behaviors, including regular COVID-19 testing, which could lead to higher detection of breakthrough infections than what is observed in the general population. Future analyses should account for testing practices when assessing breakthrough infection risk in this population. Relatedly, PWH are at higher risk for severe COVID-19 outcomes compared to PWoH.^26,27^ Because symptomatic disease is identified more frequently than asymptomatic disease, this could lead to differentially higher detection in PWH. Similarly, differentially higher detection of SARS-CoV-2 in PWH (vs. PWoH) could occur if PWH are more likely to have detectable SARS-CoV-2 virus ≥90 days after infection, as has been shown in a case report of a PWH with advanced disease and other who are immune-compromised.^28–30^ Though our matching schema was not consistent, with one cohort having already matched on demographic factors, distributions of our matching factors indicate that our sample of PWH and PWoH were comparable; we included the matching factors in multivariable analyses to address residual confounding. Lastly, observation time for individuals was necessarily short (<1 year per person) given when vaccines became available. We will continue to monitor breakthroughs monthly to accrue more follow-up time and assess breakthrough risk among PWH through December 2021. This will become especially relevant as more primary series doses and booster vaccines are administered (given recent changes in CDC recommendations) and variants, including Omicron (B.1.1.529), emerge circulate.

For PWH, the CDC recommends an additional primary series dose 28 days after the second mRNA dose, or a booster dose 2 months after a single J&J dose, among those with advanced or untreated HIV. Our findings indicate all PWH should be included in this recommendation as the risk of breakthrough was higher in PWH than PWoH regardless of CD4 count (reflecting advanced disease) or HIV viral suppression (reflecting treatment). Given our findings of breakthrough by vaccine type, clinicians should consider a booster immediately among PWH who received J&J, potentially initiating a two-dose mRNA series, as recommended by HIV Medical Association and the Infectious Diseases Society of America.^31^ The increased risk of breakthrough infections in PWH merits continued monitoring by vaccine type as the COVID-19 pandemic persists, immunity to primary vaccine series wane, boosters are widely recommended, and new variants emerge.

## Supporting information

STROBE checklist

## Data Availability

All data produced in the present study are available upon approval from the co-authors for the intended use of the data, and with data sharing approval and signed data user agremeents with each of the institutions.

