## Supplement for "COVID-19 infections post-vaccination by HIV status in the United States"

**Table S1**: Definitions of in care or in cohort, and criteria used to identify people with (PWH)

| **Cohort** | **Description** | **“In care” or “in cohort”** | **PWH** |
| --- | --- | --- | --- |
| Kaiser Permanente Mid-Atlantic States (KPMAS) | Integrated Health system | In Care – KPMAS membership for ≥1 month in 2020 | HIV registry* |
| Kaiser Permanente Northern California (KPNC) | Integrated Health system | In Care – KPNC membership for ≥1 month between 3/1/20 and 12/31/20 | HIV registry* |
| University of North Carolina Chapel Hill (UNC) | Medical center cohort | ≥1 encounter with UNC Health in 2019 and alive as of 03/01/2020 | HIV diagnosis (by ICD diagnosis code) |
| Vanderbilt University Medical Center (VUMC) | Medical center cohort | Encounter with VUMC in 2019 and alive in 2020 | HIV diagnosis code or problem list mention |
| Veterans Aging Cohort Study (VACS) | National cohort of all PWH and 1:2 demographically-matched PWoH in care in the Veterans Health Administration (VA) system | Enrolled in VACS from 1996-2017 and alive in 2020 | HIV diagnosis (presence of 1 inpatient or 2 outpatient ICD codes for HIV) |

*The Kaiser Permanente Mid-Atlantic and Northern Cali

The Kaiser Permanente Mid-Atlantic States and Northern California HIV Registries are databases of members diagnosed with HIV since 1998 and 1980, respectively. Primary sources used to identify HIV patients are HIV-specific laboratory tests, antiretroviral therapy, hospital-based HIV diagnosis, and diagnosis confirmed by infectious disease physicians.

**Figure S1:** Schematic for identification of SARS-CoV-2 breakthrough infections

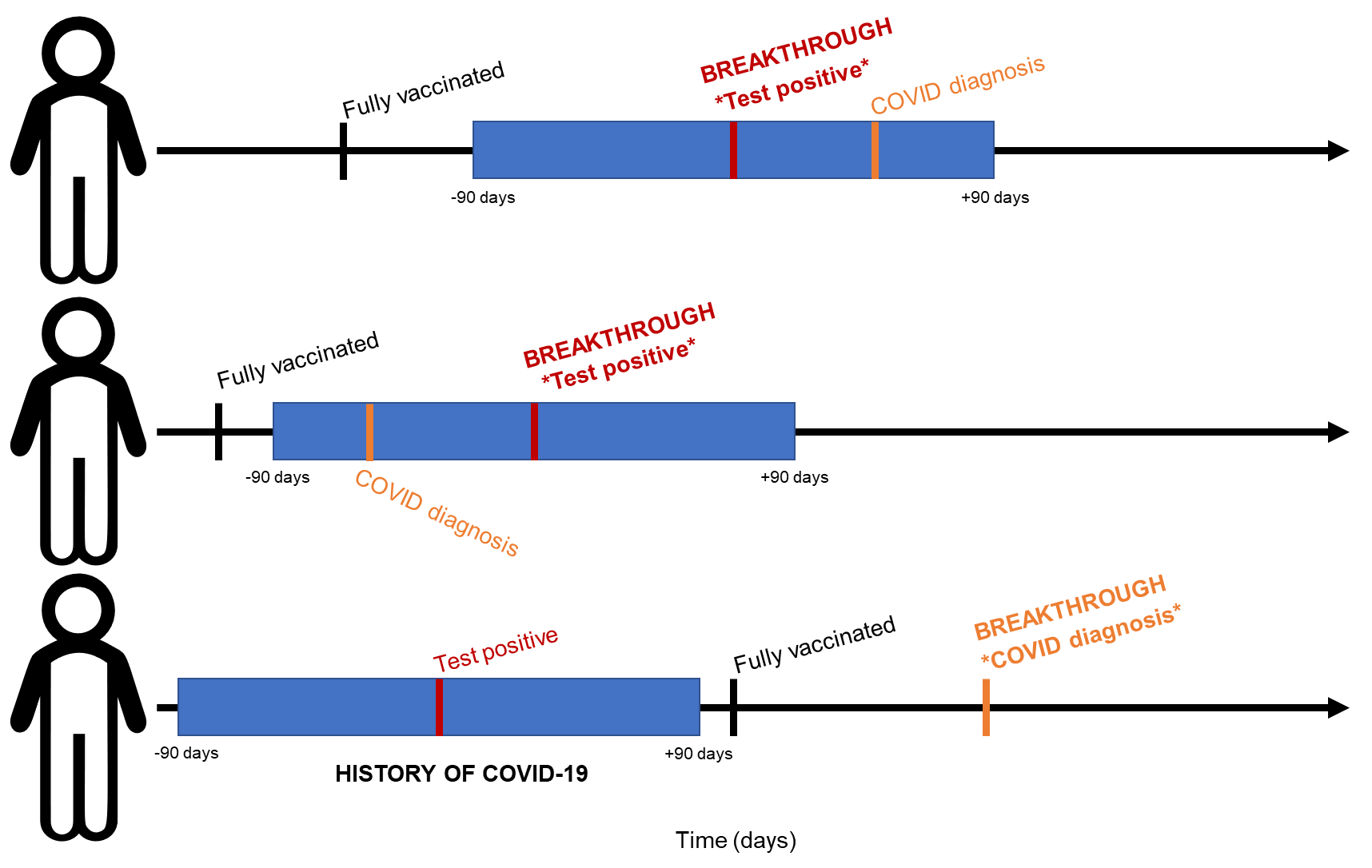

Footnotes:

Blue bar indicates +/- 90 window around a positive or detectable SARS-CoV-2 test result.

Red bar/text indicates the date of a positive or detectable SARS-CoV-2 test result.

Orange bar/text indicates the date of a COVID-19 diagnosis (using ICD-10 codes).

**Table S2:** Characteristics of PWH who did and did not receive a SARS-CoV-2 vaccine dose after their primary series (N=31,840)

| Characteristic | Overall, N = 31,840^1^ | Primary series only, N = 23,577^1^ | Received 3rd dose ≥90 days after completion of primary series, N = 8,263^1^ | p-value^2^ |
| --- | --- | --- | --- | --- |
| **Age (years)** |  |  |  | <0.001 |
| 18-24 | 81 (0.3%) | 75 (0.3%) | 6 (0.1%) |  |
| 25-34 | 1,519 (4.8%) | 1,367 (5.8%) | 152 (1.8%) |  |
| 35-44 | 3,232 (10.2%) | 2,673 (11.3%) | 559 (6.8%) |  |
| 45-54 | 5,516 (17.3%) | 4,290 (18.2%) | 1,226 (14.8%) |  |
| 55-64 | 10,622 (33.4%) | 7,978 (33.8%) | 2,644 (32.0%) |  |
| 65-74 | 8,350 (26.2%) | 5,568 (23.6%) | 2,782 (33.7%) |  |
| 75+ | 2,520 (7.9%) | 1,626 (6.9%) | 894 (10.8%) |  |
| **Sex** |  |  |  | <0.001 |
| Male | 29,636 (93.1%) | 21,790 (92.4%) | 7,846 (95.0%) |  |
| Female | 2,204 (6.9%) | 1,787 (7.6%) | 417 (5.0%) |  |
| **Ethnicity and Race** |  |  |  | <0.001 |
| Non-Hispanic White | 12,631 (39.7%) | 8,846 (37.5%) | 3,785 (45.8%) |  |
| Non-Hispanic Black/African American | 12,852 (40.4%) | 10,031 (42.5%) | 2,821 (34.1%) |  |
| Hispanic | 3,998 (12.6%) | 2,937 (12.5%) | 1,061 (12.8%) |  |
| Non-Hispanic Asian | 1,020 (3.2%) | 735 (3.1%) | 285 (3.4%) |  |
| Other | 1,040 (3.3%) | 783 (3.3%) | 257 (3.1%) |  |
| Unknown | 299 (0.9%) | 245 (1.0%) | 54 (0.7%) |  |
| **Month fully vaccinated** |  |  |  | <0.001 |
| January 2021 | 343 (1.1%) | 194 (0.8%) | 149 (1.8%) |  |
| February 2021 | 3,640 (11.4%) | 2,202 (9.3%) | 1,438 (17.4%) |  |
| March 2021 | 8,482 (26.6%) | 5,648 (24.0%) | 2,834 (34.3%) |  |
| April 2021 | 11,910 (37.4%) | 8,892 (37.7%) | 3,018 (36.5%) |  |
| May 2021 | 5,505 (17.3%) | 4,746 (20.1%) | 759 (9.2%) |  |
| June 2021 | 1,960 (6.2%) | 1,895 (8.0%) | 65 (0.8%) |  |
| **Primary vaccination series type** |  |  |  | <0.001 |
| Pfizer | 16,720 (52.5%) | 11,154 (47.3%) | 5,566 (67.4%) |  |
| Moderna | 13,174 (41.4%) | 10,530 (44.7%) | 2,644 (32.0%) |  |
| J&J | 1,946 (6.1%) | 1,893 (8.0%) | 53 (0.6%) |  |
| **COVID prior to fully vaccinated** | 2,146 (6.7%) | 1,637 (6.9%) | 509 (6.2%) | 0.015 |
| **CD4 at ART initiation (cells/mm^3^)** | 366.00 (200.00, 580.00) | 370.00 (205.00, 586.25) | 350.00 (183.00, 560.75) | <0.001 |
| Unknown | 14,530 | 10,409 | 4,121 |  |
| **AIDS before fully vaccinated** | 1,644 (14.8%) | 1,091 (14.2%) | 553 (16.2%) | 0.008 |
| Unknown | 20,738 | 15,899 | 4,839 |  |
| **CD4 at fully vaccinated (cells/mm^3^)** | 622.00 (424.00, 846.00) | 625.00 (423.00, 849.00) | 612.00 (426.00, 839.00) | 0.3 |
| Unknown | 6,090 | 4,795 | 1,295 |  |
| **Suppressed HIV RNA at fully vaccinated (<50 copies/mL)** | 24,500 (88.1%) | 17,665 (87.4%) | 6,835 (89.9%) | <0.001 |
| Unknown | 4,031 | 3,371 | 660 |  |
| ^1^n (%); Median (IQR) | | | | |
| ^2^Pearson's Chi-squared test; Wilcoxon rank sum test | | | | |

**Table S3:** SARS-CoV-2 breakthrough infection incidence rates and 95% confidence intervals, by vaccine type and HIV status

|  | **# of breakthroughs** | **# of person-years** | **IR (95% CI) per 1,000 person years** |
| --- | --- | --- | --- |
| **Moderna** |  |  |  |
| Overall | 555 | 21,663.0 | 25.6 (23.5, 28.0) |
| PWH | 192 | 6,123.1 | 31.4 (27.1, 36.1) |
| PWoH | 363 | 15,539.9 | 23.4 (21.0, 25.9) |
| **Pfizer** |  |  |  |
| Overall | 1,028 | 25,966.1 | 40.0 (37.2, 42.1) |
| PWH | 404 | 7,802.9 | 51.8 (46.9, 57.1) |
| PWoH | 624 | 18,163.2 | 34.4 (31.7, 37.2) |
| **J&J** |  |  |  |
| Overall | 184 | 3,101.6 | 59.3 (51.1, 68.5) |
| PWH | 60 | 833.2 | 72.0 (55.0, 92.7) |
| PWoH | 124 | 2,268.4 | 54.7 (45.5, 65.2) |
